# Inverted covariate effects for mutated 2^nd^ vs 1^st^ wave Covid-19: high temperature spread biased for young

**DOI:** 10.1101/2020.07.12.20151878

**Authors:** Hervé Seligmann, Siham Iggui, Mustapha Rachdi, Nicolas Vuillerme, Jacques Demongeot

## Abstract

**Background:** Here, we characterize COVID-19 2^nd^ waves, following a study presenting negative associations between 1^st^ wave COVID-19 spread parameters and temperature;

**Methods:** Visual examinations of daily increase in confirmed COVID-19 cases in 124 countries, determined 1^st^ and 2^nd^waves in 28 countries;

**Results:** 1^st^ wave spread rate increases with country mean elevation, temperature, time since wave onset, and median age. Spread rates decrease above 1000m, indicating high UV decrease spread rate. For 2nd waves, associations are opposite: viruses adapted to high temperature and to infect young populations. Earliest 2^nd^ waves started April 5-7 at mutagenic high elevations (Armenia, Algeria). 2^nd^ waves occurred also at warm-to-cold season transition (Argentina, Chile). Spread decreases in most (77%) countries. Death-to-total case ratios decrease during the 2^nd^wave, also when comparing with the same period for countries where the 1^st^ wave is ongoing. In countries with late 1^st^ wave onset, spread rates fit better 2^nd^ than 1^st^ wave-temperature patterns; In countries with ageing populations (examples: Japan, Sweden, Ukraine), 2^nd^ waves only adapted to spread at higher temperatures, not to infect children.

**Conclusions:** 1^st^ wave viruses evolved towards lower spread and mortality. 2^nd^ wave mutant COVID-19 strain(s) adapted to higher temperature, infecting children and replace (also in cold conditions) 1^st^ wave COVID-19 strains. Counterintuitively, low spread strains replace high spread strains, rendering prognostics and extrapolations uncertain.

## 1. Introduction

Spread parameters of the Covid-19 pandemics decrease with temperature [1]. This could be a direct effect of temperature causing faster aerosol evaporation, limiting travel time and distance of airborn droplets with viral particles. Alternatively, high temperature due to insulation is a proxy for ultra-violet light (UV) exposure. UVs are highly mutagenic and might decrease viral “viability”.

## 2. Methods

We follow the same methodology as in [1] by using coefficients (slopes) from regression analyses, adjusting an exponential model y = a*exp(b*x) where y is the daily number of new confirmed COVID-19 cases, x is the number of days since wave onset, a is a constant and b is the slope. This corresponds to the log-transformed version ln y = ln a + b*x. Daily numbers of new cases and deaths per countries are from [2], data on mean elevation from [3], mean temperature from [4] and counts of mutations from [5, 6].

## 3. Results

### 3.1. Relationship between Covid-19 cases and mean elevation

Figure 1 plots slopes of exponential regressions on time of daily new cases (calculated as a function of days since first 100 confirmed cases) as a function of mean country elevation. Exponents increase with temperature up to 900-1000m, then drop above 1000m, especially for landlocked high elevation countries. This analysis potentially disentangles co-linearities between temperature and UV.

**Figure 1.**
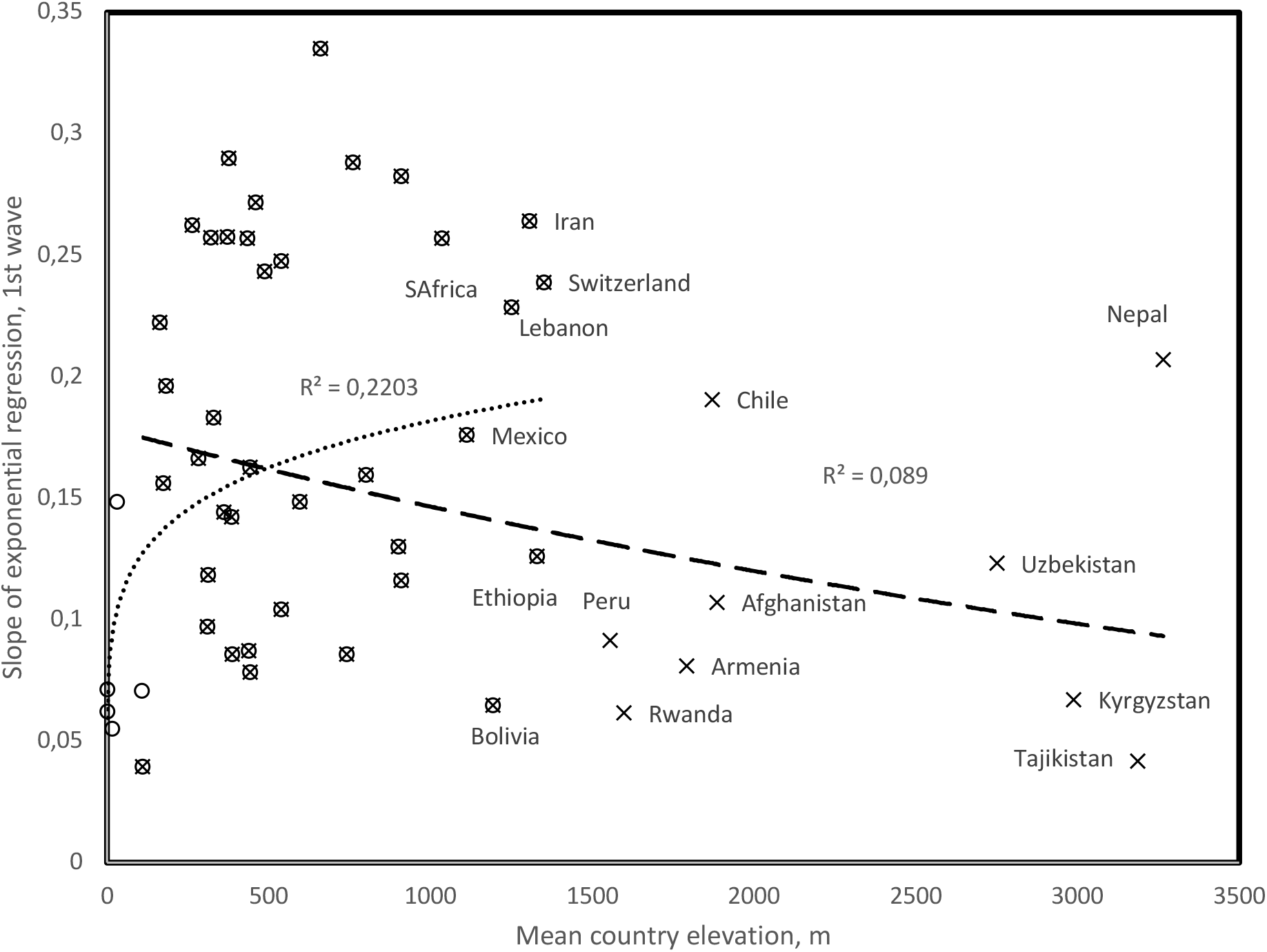
Slope of daily new confirmed Covid-19 cases as a function of mean country elevation. Circles: Countries contributing to the positive trend with elevation, up to 1400m, r = 0.469, two tailed P = 0.0015; X: Countries contributing to the negative trend with elevation, down to 110m, r = −0.298, two tailed P = 0.0417. Note that for countries above 1000m, landlocked and/or isolated countries tend to fit the negative trend (examples: Bolivia, Ethiopia, Armenia, Afghanistan) as opposed to countries with large coastal populations (examples: Chile, South Africa) and landlocked Nepal and Switzerland probably contaminated by tourists from low elevation countries. Peru has a low slope despite large coastal populations.

The trend below 1000m confirms previously described effects of temperature on spread parameters, as temperature decreases with elevation. The drop in exponents above 1000m elevation indicates direct UV effects, probably by increasing deleterious mutations. These observations are for exponents estimated for the 1^st^ Covid-19 wave, for each country (Table 1).

**Table 1.** Exponential slopes of 1^st^ and 2^nd^ Covid-19 waves in countries with two detected waves. Columns: 1. T, mean annual temperatures from [4], 2. E, mean elevation from [3], 3. D, densities from [7], 4. A, median age in that country from [8]. Start for 1^st^ wave is the date when cumulated total confirmed cases reached 100, start for 2^nd^wave is visually estimated as in Figure 3. Slopes are the exponent b from the exponential regression y = a*exp(b*x), where y is the number of new daily cases and x the number of days since 100 cumulated cases for the 1st wave, or 2nd wave start. Total numbers of cases are given for 1^st^ wave until 2^nd^ wave, and for 2^nd^ wave until 31/5 for countries with a 2^nd^ wave up to 31/5, or before 15/4 included, and after 15/4 excluded until 31/5. %Dead: Percent dead among cases for the respective periods. 1^st^ wave data were completed by data from [1] and countries with mean elevation >900m. In Kenya and Sri Lanka, erratic data prevent 1^st^ wave slope estimation.

| Country | T | E | D | A | 1<sup>st</sup> w<br>Start | S | Tot | %Dead | 2<sup>nd</sup> w<br>Start | S | Tot | %Dead |
| --- | --- | --- | --- | --- | --- | --- | --- | --- | --- | --- | --- | --- |
| Africa |  |  |  |  |  |  |  |  |  |  |  |  |
| Algeria | 22.5 | 800 | 18 | 28.1 | 20/3 | 0.1594 | 2160 | 15.56 | 07/4 | 0.0316 | 7234 | 4.38 |
| Kenya | 24.75 | 752 | 82 | 19.7 |  |  | 225 | 4.44 | 12/5 | 0.0740 | 1737 | 3.11 |
| Ethiopia* | 22.2 | 1330 | 101 | 17.9 | 21/4 | 0.1259 | 85 | 3.53 |  |  | 1087 | 0.74 |
| Morocco* | 17.1 | 909 | 80 | 29.3 | 21/3 | 0.1161 | 2024 | 6.28 |  |  | 5783 | 1.35 |
| Rwanda* | 17.85 | 1598 | 470 | 19.0 | 04/4 | 0.0615 | 136 | 0 |  |  | 234 | 0.43 |
| South Africa* | 17.75 | 1034 | 48 | 27.1 | 17/3 | 0.257 | 2506 | 1.36 |  |  | 30177 | 2.15 |
| Asia |  |  |  |  |  |  |  |  |  |  |  |  |
| Afghanistan* | 12.6 | 1885 | 49 | 18.9 | 26/3 | 0.107 | 784 | 3.19 |  |  | 14421 | 1.61 |
| Bahrain [1] | 27.15 | 1 | 1983 | 32.3 | 09/3 | 0.1884 | 1671 | 0.42 |  |  | 9727 | 0.12 |
| Iran | 17.25 | 1305 | 51 | 30.3 | 26/2 | 0.2641 | 94640 | 6.37 | 01/5 | 0.0438 | 32309 | 3.58 |
| Iraq | 14.03 | 312 | 90 | 20.0 | 14/3 | 0.1184 | 1400 | 6.07 | 15/4 | 0.041 | 2324 | 2.41 |
| Japan [1] | 11.15 | 438 | 333 | 47.3 | 20/2 | 0.0872 | 8626 | 2.06 |  |  | 8225 | 8.67 |
| Kazakhstan | 6.4 | 387 | 7 | 30.6 | 26/3 | 0.0856 | 4834 | 0.56 | 08/5 | 0.0933 | 3488 | 0.12 |
| Kyrgyzstan | 1.55 | 2988 | 32 | 26.5 | 30/3 | 0.0671 | 656 | 1.07 | 25/4 | 0.0271 | 747 | 0.8 |
| Lebanon | 16.4 | 1250 | 672 | 30.5 | 14/3 | 0.2286 | 678 | 3.1 | 19/4 | 0.0757 | 437 | 1.14 |
| Malaysia [1] | 25.4 | 538 | 99 | 28.5 | 08/3 | 0.1042 | 6704 | 1.52 | 12/5 | 0.0794 | 519 | 1.16 |
| Nepal* | 8.1 | 3265 | 201 | 24.1 | 06/5 | 0.207 | 15 | 0 |  |  | 1557 | 0.51 |
| Oman | 25.6 | 310 | 15 | 25.6 | 25/3 | 0.0972 | 2447 | 0.45 | 02/5 | 0.0936 | 5323 | 0.47 |
| Pakistan* | 20.20 | 900 | 274 | 23.8 | 15/3 | 0.1301 | 6383 | 1.74 |  |  | 60074 | 2.28 |
| Philippines | 25.85 | 442 | 362 | 23.5 | 14/3 | 0.1627 | 5453 | 0.62 | 22/5 | 0.1772 | 12633 | 7.31 |
| Singapore [1] | 26.45 | 15 | 7894 | 34.6 | 28/2 | 0.0551 | 3699 | 0.27 | 02/5 | 0.0641 | 31189 | 0.04 |
| South Korea | 11.5 | 282 | 517 | 41.8 | 20/2 | 0.1664 | 10591 | 2.12 | 06/5 | 0.0585 | 877 | 5.13 |
| Sri Lanka | 26.95 | 228 | 332 | 32.8 |  |  | 238 | 2.94 | 08/5 | 0.1347 | 1395 | 0.22 |
| Tajikistan* | 2.00 | 3186 | 64 | 24.5 | 02/5 | 0.0418 | 0 |  |  |  | 3930 | 1.2 |
| Uzbekistan | 12.05 | 2750 | 73 | 28.6 | 28/3 | 0.1231 | 1862 | 0.38 | 26/4 | 0.0238 | 1077 | 0.46 |
| Australia [1] | 21.65 | 330 | 3 | 38.7 | 09/3 | 0.1832 | 6447 | 0.98 |  |  | 478 | 5.35 |
| Europe |  |  |  |  |  |  |  |  |  |  |  |  |
| Armenia | 7.15 | 1792 | 99 | 35.1 | 18/3 | 0.0809 | 1111 | 1.53 | 05/4 | 0.057 | 8171 | 1.4 |
| Austria [1] | 6.35 | 910 | 106 | 44.0 | 08/3 | 0.2825 | 14350 | 2.74 |  |  | 2381 | 11.55 |
| Azerbaijan | 11.95 | 384 | 116 | 32.3 | 25/3 | 0.1422 | 1592 | 1.32 | 25/4 | 0.0676 | 2039 | 1.08 |
| Belgium [1] | 9.55 | 181 | 378 | 41.4 | 06/3 | 0.1963 | 44636 | 15.48 |  |  | 10658 | 20.95 |
| Czech Rep. [1] | 7.55 | 433 | 135 | 42.1 | 11/3 | 0.257 | 6301 | 2.63 | 13/5 | 0.0474 | 2967 | 5.19 |
| Denmark |  |  |  | 42.2 |  |  | 7609 | 3.77 |  |  | 2672 | 5.31 |
| France [1] | 10.7 | 375 | 123 | 41.4 | 29/2 | 0.2898 | 161476 | 14.01 |  |  | 20087 | 27.47 |
| Germany [1] | 8.5 | 263 | 233 | 47.1 | 29/2 | 0.2624 | 151819 | 3.86 |  |  | 21926 | 10.91 |
| Italy [1] | 12.45 | 538 | 200 | 45.5 | 22/2 | 0.2475 | 174194 | 14.32 |  |  | 32013 | 18.57 |
| Lithuania | 6.2 | 110 | 73 | 43.7 | 21/3 | 0.0394 | 1418 | 2.61 | 05/5 | 0.0554 | 204 | 8.82 |
| Malta | 19.2 | 1 | 1567 | 41.8 | 23/3 | 0.0712 | 426 | 0.47 | 19/4 | 0.0536 | 184 | 1.63 |
| N Macedonia | 9.8 | 741 | 81 | 37.9 | 21/3 | 0.0858 | 1506 | 5.45 | 03/5 | 0.0528 | 472 | 6.57 |
| Netherlands [1] | 9.25 | 30 | 421 | 42.6 | 05/3 | 0.2485 | 28153 | 11.13 |  |  | 5956 | 15.43 |
| Norway [1] | 1.5 | 460 | 17 | 39.2 | 09/3 | 0.2716 | 6384 | 4.9 |  |  | 788 | 5.71 |
| Poland | 7.85 | 173 | 123 | 40.7 | 14/3 | 0.1562 | 7582 | 3.77 | 05/4 | 0.0094 | 16204 | 4.8 |
| Portugal | 15.15 | 372 | 112 | 42.2 | 13/3 | 0.0301 | 27268 | 4.05 | 09/5 | 0.0431 | 3355 | 5.87 |
| Spain [1] | 13.3 | 660 | 93 | 42.7 | 25/2 | 0.335 | 217368 | 10.44 |  |  | 55765 | 8.94 |
| Sweden [1] | 2.1 | 320 | 23 | 41.2 | 05/3 | 0.2572 | 17216 | 12.66 |  |  | 13346 | 12.28 |
| Switzerland [1] | 5.5 | 1350 | 208 | 42.4 | 04/3 | 0.2388 | 26336 | 4.71 |  |  | 4526 | 15.05 |
| UK [1] | 8.45 | 162 | 280 | 40.5 | 04/3 | 0.2223 | 14915 | 15.15 |  |  | 23574 | 13.37 |
| North America |  |  |  |  |  |  |  |  |  |  |  |  |
| Canada [1] | -5.35 | 487 | 4 | 42.2 | 10/3 | 0.2432 | 28379 | 3.56 |  |  | 62568 | 10.05 |
| Cuba | 25.2 | 108 | 102 | 41.5 | 27/3 | 0.0706 | 814 | 2.95 | 17/5 | 0.0517 | 1231 | 4.79 |
| El Salvador | 24.45 | 442 | 319 | 27.1 | 10/4 | 0.0783 | 159 | 3.77 | 21/4 | 0.0535 | 2358 | 1.7 |
| Guatemala | 23.56 | 759 | 162 | 22.1 | 09/3 | 0.088 | 180 | 2.78 | 01/5 | 0.1109 | 4559 | 2.13 |
| Panama | 25.4 | 360 | 56 | 29.2 | 18/3 | 0.1443 | 3751 | 2.75 | 19/5 | 0.1195 | 9712 | 2.4 |
| Mexico* | 21.00 | 1111 | 64 | 28.3 | 18/3 | 0.1759 | 5399 | 7.52 |  |  | 82113 | 11.41 |
| USA [1] | 8.55 | 760 | 34 | 38.1 | 02/3 | 0.2882 | 95377 | 5.68 |  |  | 623445 | 6.53 |
| South America |  |  |  |  |  |  |  |  |  |  |  |  |
| Argentina | 14.8 | 595 | 16 | 31.7 | 18/3 | 0.1485 | 4783 | 5.27 | 05/5 | 0.0427 | 4026 | 3.55 |
| Bolivia* | 21.55 | 1192 | 10 | 24.3 | 30/3 | 0.0647 | 397 | 7.05 |  |  | 9197 | 3.07 |
| Brazil |  |  |  | 32.6 |  |  | 58465 | 6.92 |  |  | 216766 | 6.85 |
| Chile | 8.45 | 1871 | 23 | 34.4 | 15/3 | 0.1906 | 14885 | 1.42 | 30/4 | 0.0586 | 38732 | 0.85 |
| Peru* | 19.6 | 1555 | 25 | 28.0 | 29/3 | 0.0915 | 11475 | 2.21 |  |  | 153001 | 2.78 |

### 3.2. Covid-19 viruses evolve over time

Numbers of mutations in a country increase with time since 1^st^ wave onset (r = 0.561, two-tailed P = 0.00084, Figure 2). Time since onset is indeed proportional to replicational cycles, and viral population evolution. No meaningful correlation was observed between mutation numbers and country mean temperature or elevation. Results remain qualitatively unchanged after excluding from analysis extreme datapoints (Nepal, UK).

**Figure 2.**
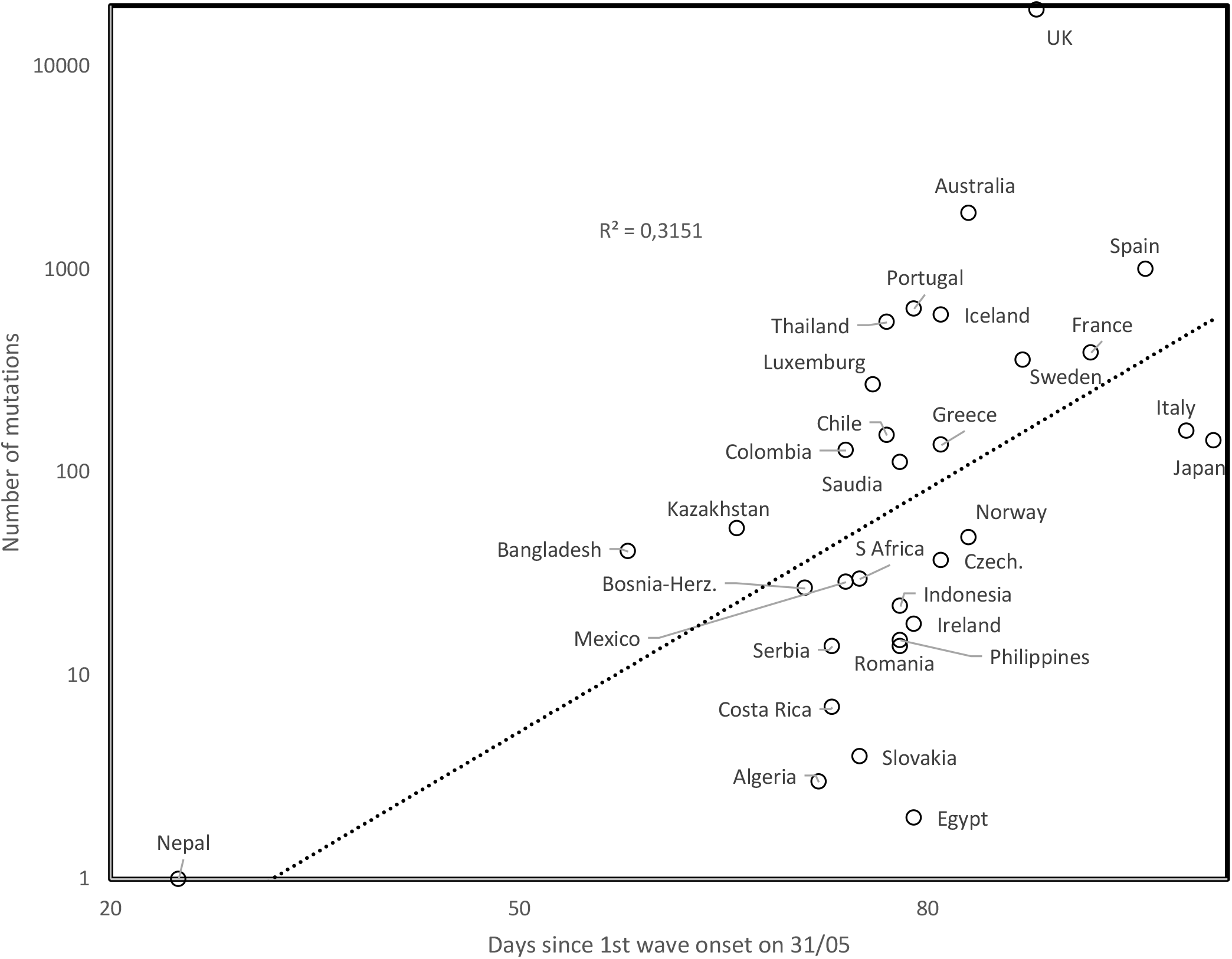
Mutation numbers as a function of days since onset of 1^st^ wave (determined on 31/05).

### 3.3. Determination of 1^st^ and 2^nd^ waves

Here, we study exponents estimated for 2^nd^ Covid-19 waves, derived from visually examining daily new cases in 123 countries. We explore temporal-, geographic-, demographic- and temperature-associated patterns of 2^nd^ wave spread parameters. We examined graphs plotting daily numbers of new confirmed cases (as daily updated at https://www.worldometers.info/coronavirus/ [2]) for 123 countries. 2^nd^ waves were visually determined, with examples in Figure 3 (Iran, Argentina). 2^nd^waves occur in 26 countries, along patterns shown for Iran (broken 1^st^ wave, 2^nd^ wave starts from a low rates). The pattern shown for Argentina (new slope after inflection in 1^st^ wave still in its growing phase) occurs also only in Chile. For Argentina and Chile, the new 2^nd^ wave slope occurred during hot-to-cold season transitions, in early April, corresponding to early October northern hemisphere seasonal shifts.

**Figure 3.**
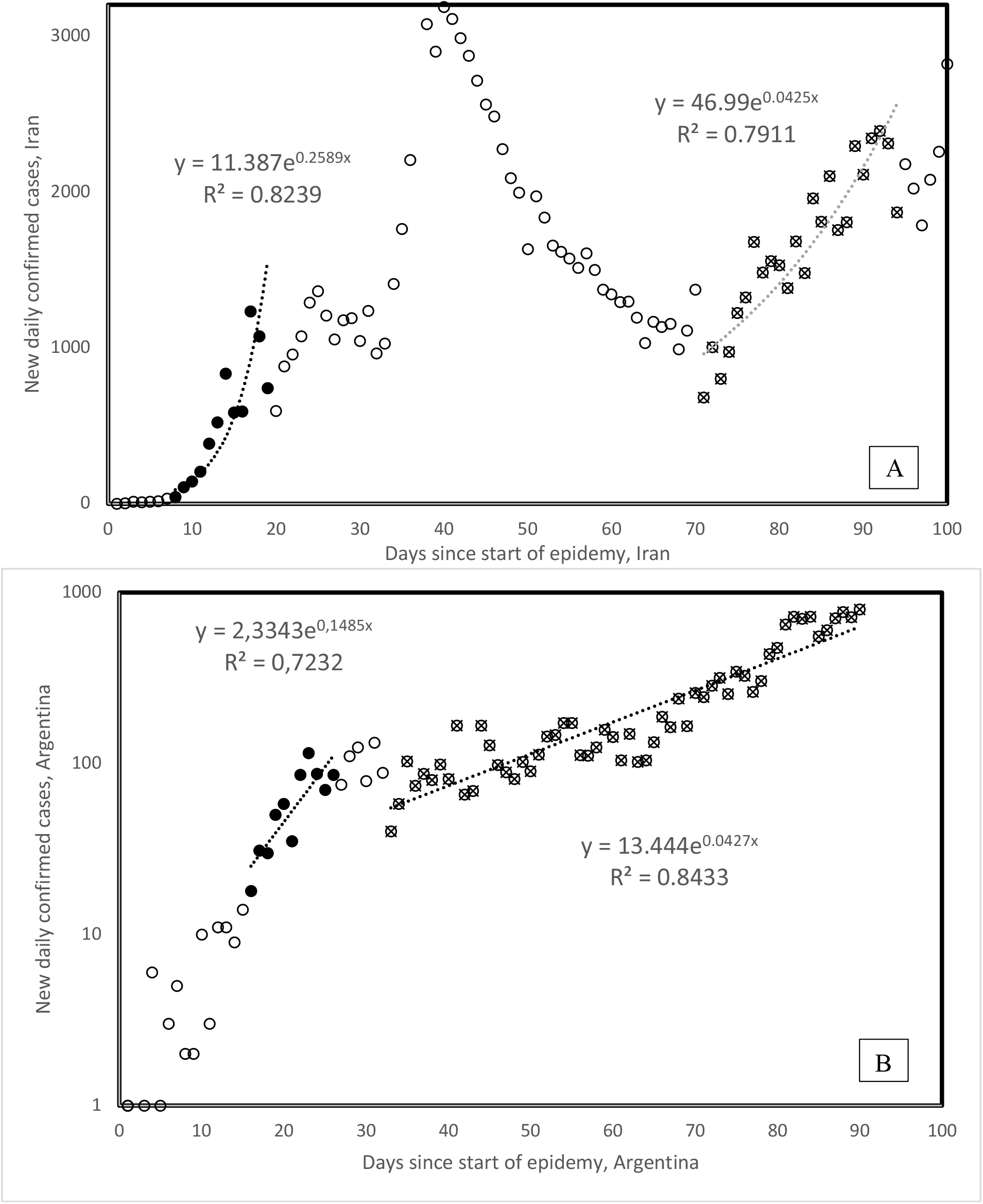
1^st^ and 2^nd^ waves of Covid-19 epidemy in Iran (A) and Argentina (B). 1^st^ wave onsets are defined from the day the cumulative total number of confirmed cases passes 100 cases. Onset of 2^nd^waves is determined visually. All countries but Chile follow the general pattern as in the example for Iran, where the new wave follows a decrease, Chile follows the pattern of Argentina. Note the log scale for the Figure 2B y axis, visually enhancing slope change.

The lower 2^nd^ vs 1^st^ wave slopes in Figure 3 are not due to temperature increase as could be expected from negative correlations between 1^st^ wave slopes and temperature [1]. This is because for Argentina and Chile (Table 1), lower slopes correspond to hot-to-cold season transition, not to cold-to-hot seasons. Table 1 compares 1^st^ and 2^nd^ wave slopes.

### 3.4. Geographical 2^nd^ wave clusters

Visual data examinations such as in Figure 3 for 123 countries detect 2^nd^ waves in 28 countries from four continents (Africa (2), America (North, 4; South, 2), Asia (12) and Europe (8)). For Kenya and Sri Lanka, 1st wave slopes could not be determined (Table 1). Earliest 2^nd^ waves are from Armenia and Poland (April 5) and Algeria (April 7). 2^nd^ waves are distributed into 4 geographic clusters (from earliest to latest): one spreading from the high elevation Eurasian plateau to surrounding countries (5/4 Armenia, 15/4 Iraq, 19/4 Lebanon, 25/4 Azerbaijan, Kyrgyzstan, 26/4 Uzbekistan, 1/5 Iran, 2/5 Oman, 8/5 Kazakhstan), a Central American cluster (21/4 El Salvador, 1/5 Guatemala, 17/5 Cuba, 19/5 Panama), a South American cluster (30/4 Chile, 5/5 Argentina), and a South-East Asian cluster (2/5 Singapore, 8/5 Sri Lanka, 12/5 Malaysia, 22/5 Philippines).

### 3.5. 2^nd^ wave slopes and mortalities

2^nd^ wave slopes are lower than 1^st^ wave slopes for 20 among 26 countries (exceptions: Guatemala, Kazakhstan, Lithuania, Philippines, Portugal, Singapore), a statistically significant majority (two tailed sign test, P = 0.0047). Mean 2^nd^ wave slope decrease as compared to 1^st^ is by 43%.

Percentages of fatal cases among all confirmed cases are lower during 2^nd^ waves than during 1^st^waves in 17 among 28 countries (not a statistically significant majority). For countries where only the 1^st^ wave occurred, mortality among confirmed cases increases in 20 among 29 countries during the 2^nd^ wave period (16/4-31/5) as compared to the period up to 15/4. This is a statistically significant majority of countries (two tailed sign test, P = 0.031). The reasons for this increase are unknown and would suggest that at the end of epidemic waves, in this case the 1^st^ wave, more lethal forms arise. Note that methods used here can not assess whether mixtures of 1^st^ and 2^nd^ wave virus strains occur in countries.

Differences between these two country groups are statistically significant (two-tailed Fisher exact test, P = 0.0343). Hence mortality rates seem to increase in 1^st^ wave countries during the period the 2^nd^wave hits other countries, where mortality slightly decreases. Overall, 2^nd^ waves are characterized by slower spread and perhaps lower mortality as compared to the same period for 1^st^waves.

### 3.6. 2^nd^ wave spread rates and temperature

Figure 4 plots 1^st^ and 2^nd^ wave slopes as a function of mean annual temperature. Analyses for the 16 countries from [1] show a negative association between 1^st^ wave slope and mean annual temperature (open circles in Figure 4), producing r = −0.606, two tailed P = 0.0128. The overall pattern for the 1^st^ wave (37 countries added, filled circles and triangles) remains qualitatively as in [1] (r = −0.329, P = 0.00817, one tailed test, considering all 53 countries).

**Figure 4.**
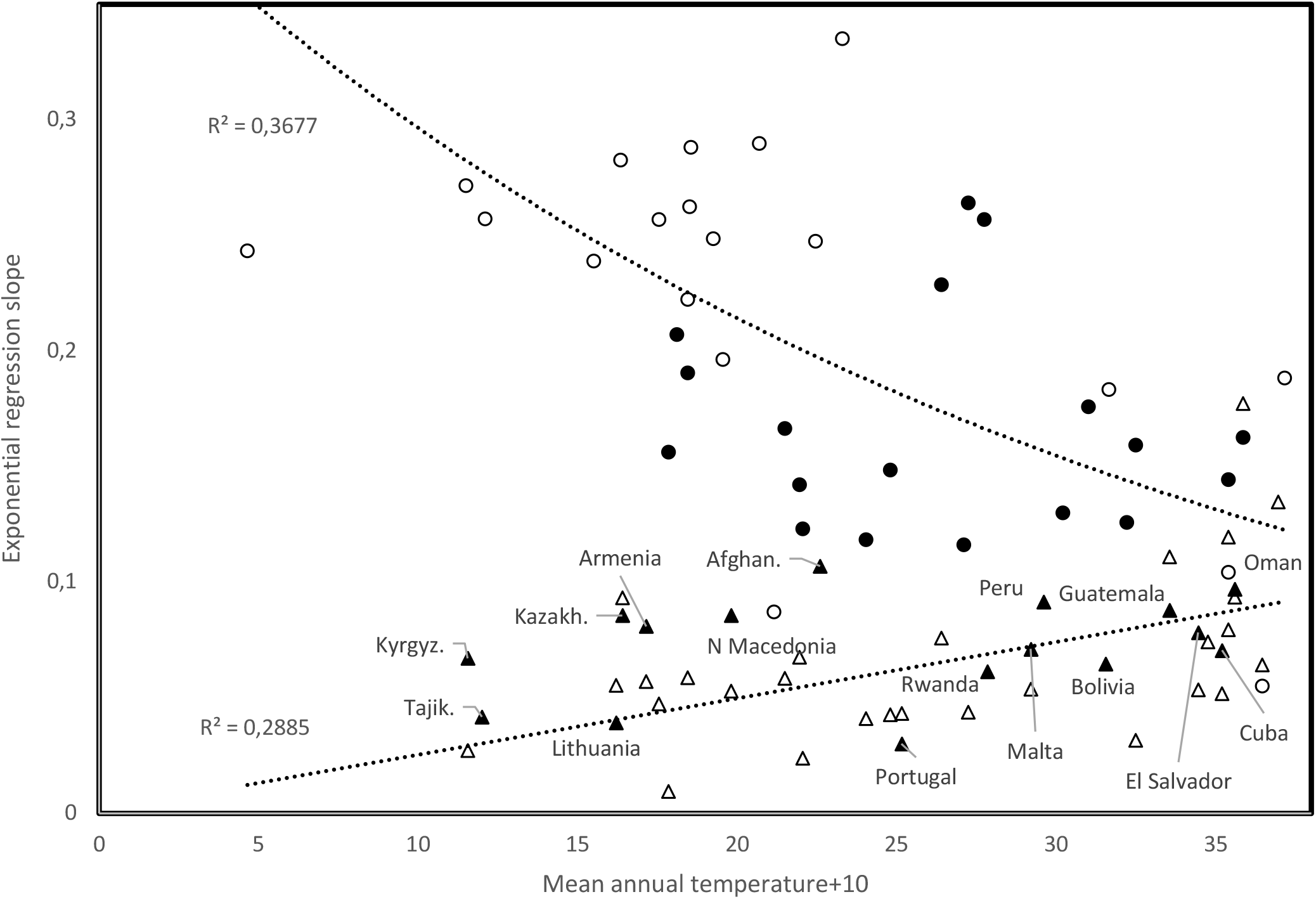
Slope of exponential regression of daily new cases vs time, as a function of mean annual temperature, comparing trends for 1^st^ wave slopes (open circles from [1], filled circles and triangles are from this study), and 2^nd^ wave slopes (open triangles).

2^nd^ wave slopes (open triangles, Figure 4) increase with temperature (r = 0.537, two tailed P = 0.00321). Unknown mechanisms enable 2^nd^ wave viral populations spread at high temperatures. Earliest 2^nd^ wave occurrences at high elevations (Armenia, Algeria) might not be circumstantial: high UV regimes, increasing mutation rates, might occasionally favour selection of temperature-adapted viruses.

### 3.7. Time since start of 1^st^ wave for low slopes

Filled triangles in Figure 4 are for countries whose 1^st^ wave slope fits better the increasing 2^nd^wave trend than the decreasing 1^st^ wave trend (2^nd^ wave onset date before country): 9/3 Guatemala, 13/3 Portugal, 18/3 Armenia, 21/3 Lithuania, North Macedonia, 23/3 Malta, 25/3 Oman, 26/3 Afghanistan, Kazakhstan, 27/3 Cuba, 29/3 Peru, 30/3 Bolivia, Kyrgyzstan, 4/4 Rwanda, 10/4 El Salvador, 2/5 Tajikistan. On 31/5, the mean time since 1^st^ wave onset in these countries was 65.19 days, significantly less than 76.32 days since 1^st^ wave onset in remaining countries that fit best the negative trend (two tailed t-test, P = 0.0228). Hence, 1^st^ wave viral population dynamics evolved towards low spread.

### 3.8. Slopes and times since start of 1^st^ and 2^nd^ waves

Time since 1^st^ wave onset increases with spread slope (r = 0.4968, P = 0.00018, two tailed test, Figure 5a). More recent 1st waves have lower mortality rates (r = 0.4194, P = 0.002, two tailed test). Outliers with high slopes despite recent start associate with high elevation, outliers with low slope despite early 1^st^wave have developed marine commerce. Note that this trend observed across countries does not contradict results described above for specific countries where mortalities increase as compared to the first period of the 1^st^ wave in that country. Time since 1^st^ wave start could be a proxy of temperature, as early 1^st^ waves occurred in February vs late ones in April. Seasonal temperature might decrease slopes at their start for countries that joined late the 1^st^ wave. However, mean annual temperature across countries does not correlate with the time since 1^st^ wave onset. Hence, the effect in Figure 5a seems independent from mean temperature.

**Figure 5.**
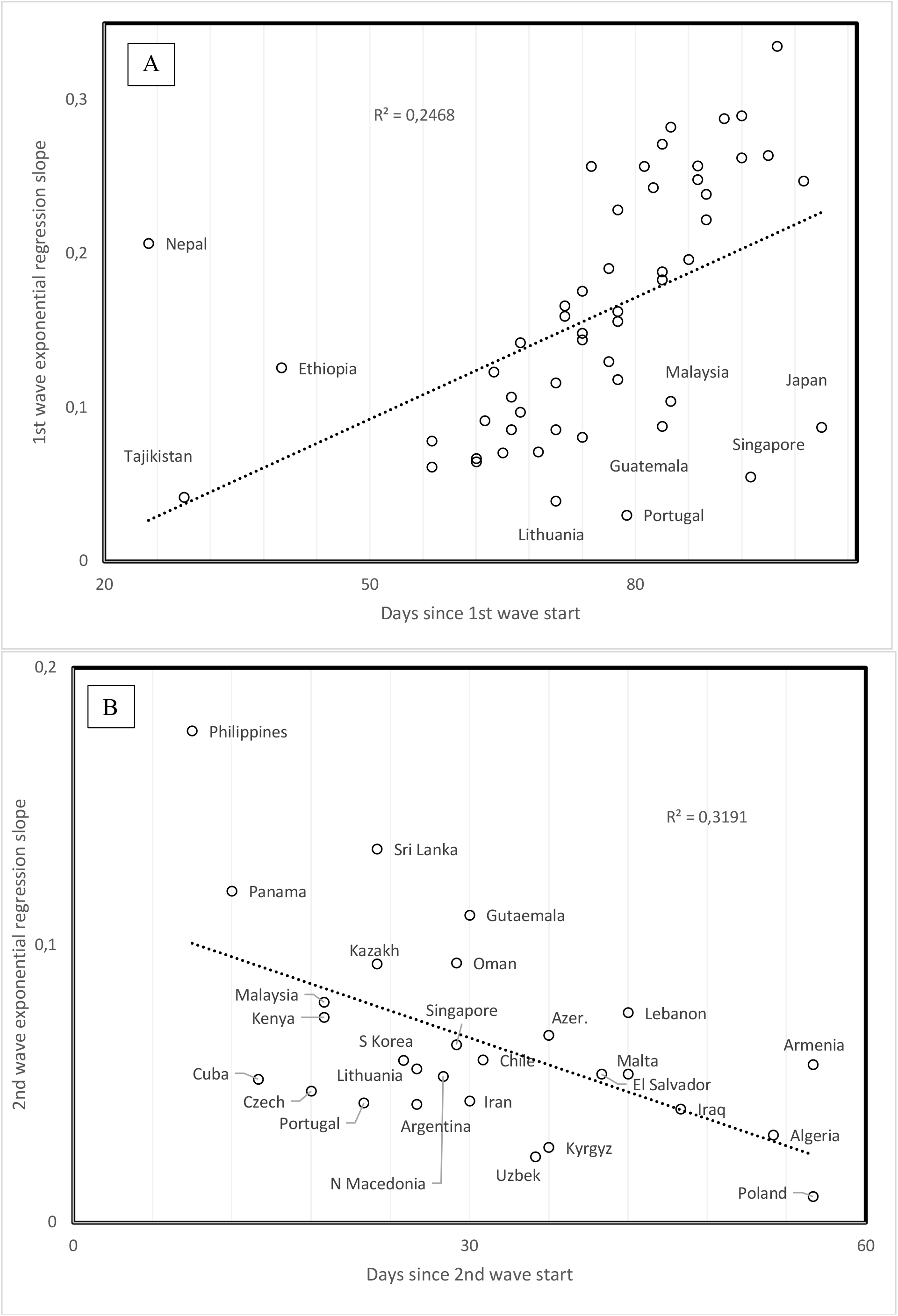
Slope of daily increase in confirmed COVID-19 cases as a function of time since wave started for A. 1^st^and B. 2^nd^ waves.

Considering that time is proportional to replication cycles of viral populations (Figure 2), results indicate that 1^st^ wave viruses evolve towards lower slopes and mortalities at the level of comparisons between countries. This again contrasts with patterns for the 2^nd^ wave, where time since 2^nd^ wave onset correlates negatively with 2^nd^ wave slope (r = −0.5649, P = 0.0026, two tailed test, Figure 5b). Mortality also tends to increase but this trend is not statistically significant (r = −0.2439, P = 0.2299, two tailed test). Hence, 2^nd^ wave viral populations might increase their spread over time, possibly implying adaptation.

### 3.9. Elevation and population density

Mean country elevation correlates negatively with time since 1^st^ wave onset (r = −0.6095, P = 0.0000016, two tailed test). This suggests that the pandemic reached later more elevated and possibly isolated countries. Low elevation also associates with ports and probable spread via marine commerce.

Notable is that no pandemic property (Table 1) correlate with population density. One would have expected that slopes increase with population densities, but this is not the case (1^st^ wave: r = −0.1779, P = 0.2068; 2^nd^ wave: r = 0.0128, P = 0.94845; two tailed tests). It seems that most COVID-19 cases are in dense urban centres. These densities might vary among different cities, but mean country does not reflect this. New York city and Singapore might have the same urban densities, but population densities for their respective countries vary due to differences in sizes of surrounding low population areas. Hence, no correlation could be observed using our simple method.

### 3.10. Median age and spread rates

1^st^ wave virus strains hit mainly the elderly. Hence the positive correlation between slope and median population age in Figure 6a (r = 0.414, one tailed P = 0.0011) fits the expected higher contagiousness in ageing populations. Note outliers as indicated in Figure 6a. For the 2^nd^ wave, the opposite association occurs: slopes are highest for countries with low median age (r = −0.418, two tailed P = 0.0023, Figure 6b). This new information is crucial for future management of the pandemic: 2^nd^ wave viruses apparently adapted to infect the larger reservoir of potential younger hosts, in addition to adapting to spread at higher temperatures.

**Figure 6.**
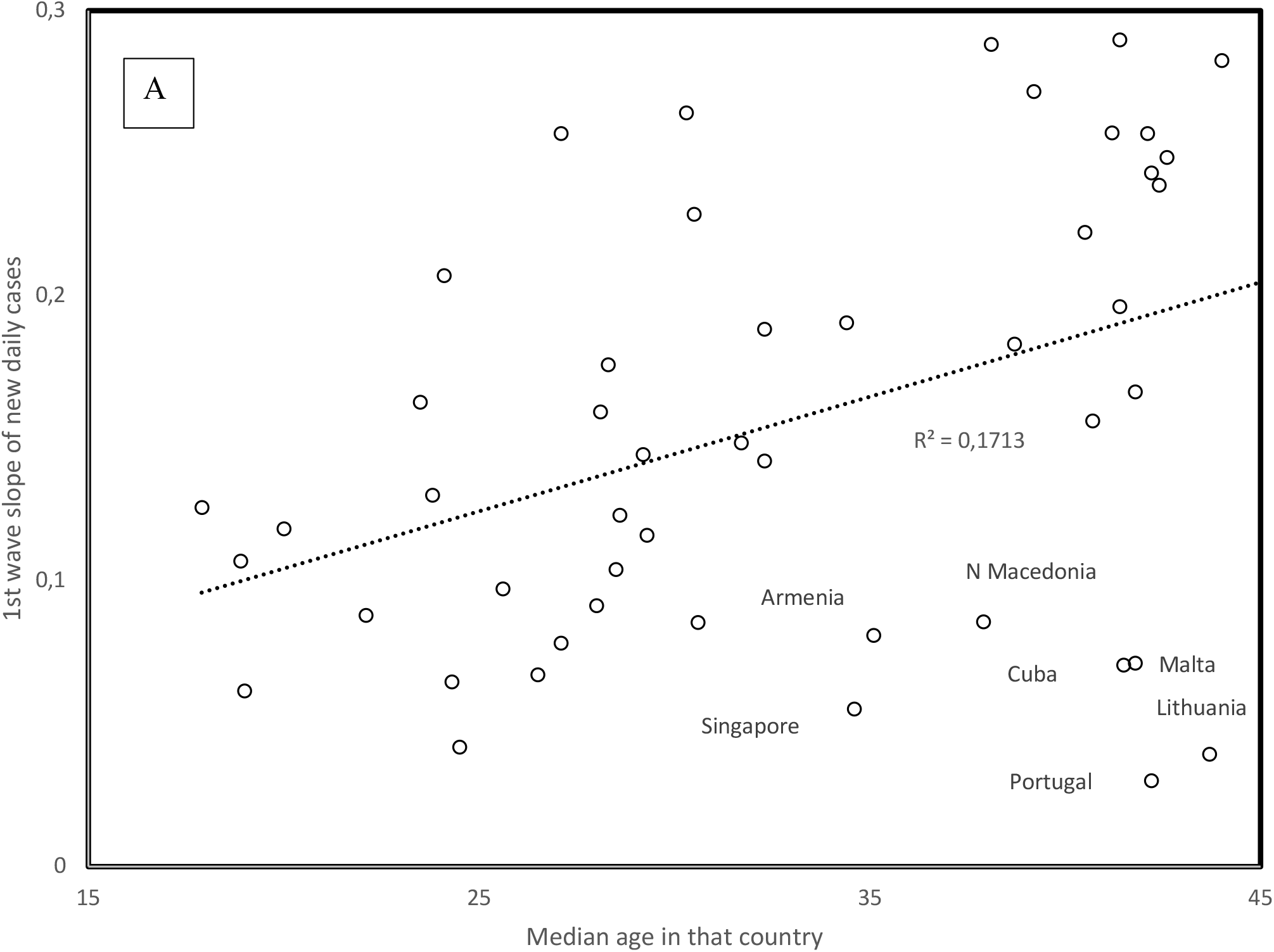
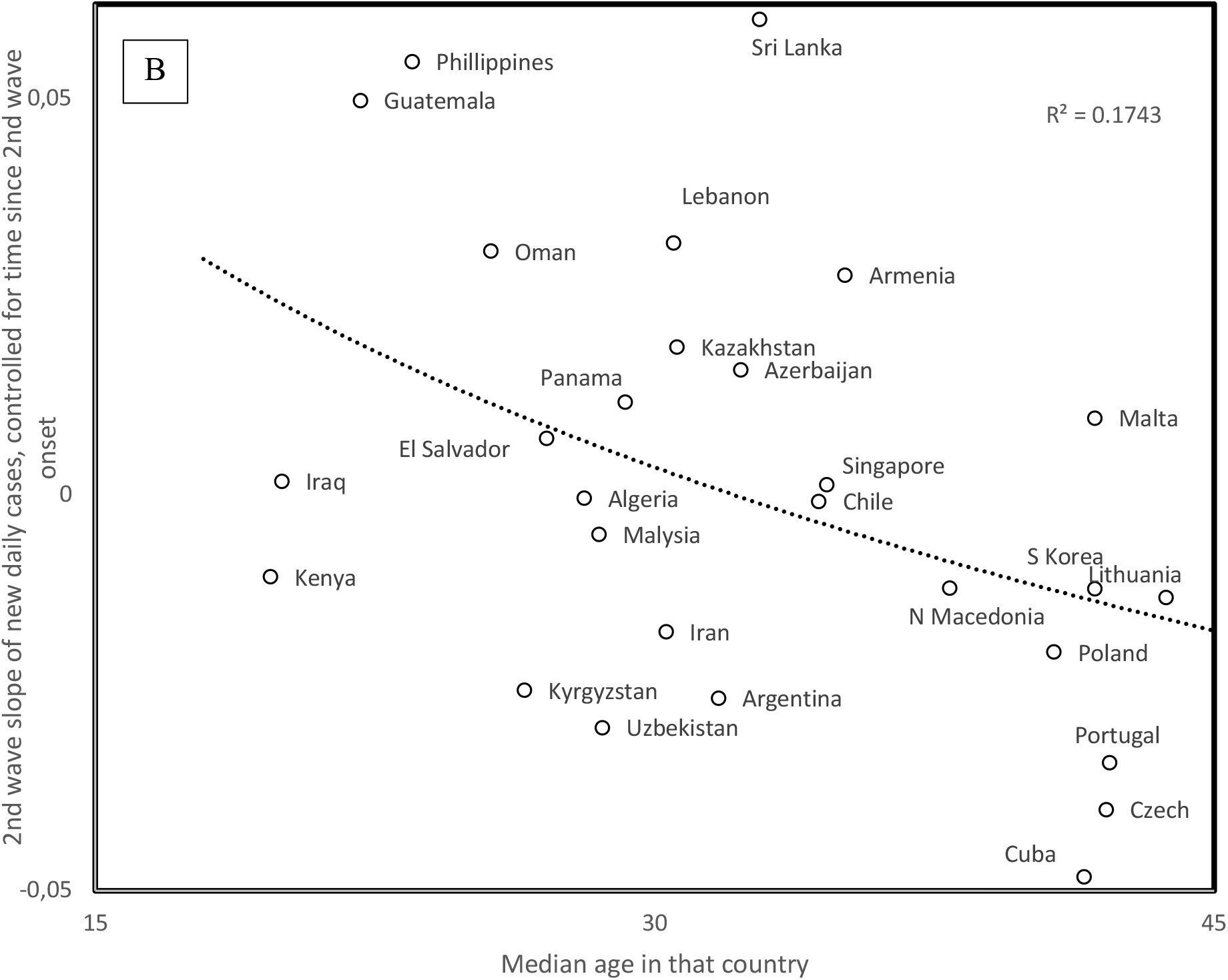
1^st^ (A) and 2^nd^ (B) wave slopes as a function of country median age. For 2^nd^ wave slopes, the figure plots the residual values after adjusting 2^nd^ wave slopes for time since 2^nd^ wave start (regression in Figure 4B), the main correlate of 2^nd^ wave slopes.

Data gathered after 31/5 find 2^nd^ waves in additional countries. For countries with median ages above 36 years (Bulgaria, Japan, Moldova, Serbia, Sweden, Ukraine), the trend for these 2^nd^ wave slopes fits that observed for the 1^st^ wave slopes as a function of median age in Figure 6a. This indicates that in these countries with late 2^nd^ wave onset and high median age, viruses only adapted to seasonal temperature increase, but not to the relatively few young in these populations. In some countries, the 2^nd^ wave might be an artefact due to sudden policy changes such as increasing daily test numbers, which increase numbers of new reported cases, but do not reflect any biological change. Other 2^nd^wave slopes estimated after 31/5 fit the trend in Figure 6b for Bahrain, Democratic Republic of Congo, Ghana, Guatemala, Iran, Israel, and Jordan. These patterns could be explained by policy differences between countries. Our interpretation remains biological, and suggests that viruses evolve in relation to host populations and climatic conditions because country-specific sampling artefacts are unlikely to produce overall patterns across countries such as those in Figures 5 and 6.

## 3. Discussion

Analyses confirm that the spread of 1^st^ wave COVID-19 decreases with temperature. They indicate that UV also decreases the spread of 1^st^ wave COVID-19. 2^nd^ wave COVID-19 is characterized by a lower spread and mortalities lower than for the comparable period for 1^st^ waves, and by infecting younger age classes. 2^nd^ wave spread increases with temperature.

This inversion of trends between 1^st^ and 2^nd^ waves, at a 1-2 months interval, is highly peculiar. The possibility that a different virus was cryptic and minor during the 1^st^ wave and became dominant as conditions changed during the 2^nd^ wave, cannot be excluded. However, trends with time and mutation numbers suggest that a specific virus evolved from one state to another. The earliest 2^nd^ waves, in high elevation countries, suggest UV-induced high mutation rates hastened adaptation.

Note that analyses determined clear patterns in relation to various cofactors of the pandemic, despite uncertainties in data. For example, reported over unreported cases ratio [10] apparently vary hugely between countries depending on their mode of counting and public health policy, rendering predictions for the future of the pandemic highly uncertain. For the 1^st^ wave, within countries, mortalities increased despite the general trend across countries for decrease of mortality over time. It is too early for exploring 2^nd^ wave-associated patterns within countries regarding the evolution of mortalities over time, and in relation to symptoms developed in the young. A 3^rd^ wave may combine 1^st^ wave high spread (and mortality) and 2^nd^ wave tolerance for high temperature. A striking major point is unexplained and open for more optimistic interpretations from a health-oriented point of view. Usually, strains with high spread replace those with low spread. However, low spread 2^nd^ wave viruses replace in an increasing number of countries fast spread 1^st^ wave viruses. Hence, accepted knowledge in relation to epidemics seems inadequate regarding the current pandemic. Hence, prognostics and interpretations of observed patterns, whether pessimistic or optimistic, cannot be trusted, as these are based on previous knowledge that contradicts the current fast-to-slow spread evolution of COVID-19.

## Data Availability

All data are available in the manuscript

## Author Contributions

Conceptualization, methodology, investigation, J.D., S.Y., H.S.; resources, J.D., S.Y., H.S.; data curation, S.Y., H.S.; writing—original draft preparation, H.S.; writing—review and editing, J.D. All authors have read and agreed to the published version of the manuscript.

